# Integrating Computational Optimization with Antimicrobial Susceptibility Testing: A Particle Swarm Optimization Framework for Enhancing Fluoride Toothpaste Formulations

**DOI:** 10.64898/2026.03.25.26349293

**Authors:** Clive Asuai, Oscar Whiliki, Andrew Mayor, Dennis Victory, Omonigho Imarah, Debekeme Irene, Ighere Merit, Houssem Hosni, Muhammad Ibrahim Khan, Akpevweoghene Courage Edwin

## Abstract

This study develops a methodological framework that combines conventional antimicrobial susceptibility testing with Particle Swarm Optimisation (PSO) to enhance toothpaste formulations, employing Escherichia coli isolated from the oral cavity as a model organism.

We used the agar well diffusion method to see if two fluoride toothpastes (Oral B and My-my) could kill oral E. coli isolates at 6.25%, 12.5%, 25%, 50%, and 100% concentrations. A surrogate Random Forest model was created using these experimental data to link formulation parameters to antimicrobial activity. Then, PSO was used to find the best formulation traits. Multi-objective optimisation that looks at the trade-offs between antimicrobial effectiveness and cytotoxicity was shown as a conceptual framework.

Both toothpastes showed antimicrobial activity that depended on the concentration, with Oral B being more effective (23.0 mm at 100% concentration) than My-my (20.0 mm). The PSO framework, utilised as a methodological illustration while explicitly recognising data constraints, determined hypothetical formulation parameters (sodium fluoride 1100 ppm, hydrated silica abrasive, 2.5% SLS) with an anticipated zone of inhibition of 26.3 mm. These predictions are mathematically optimal for a surrogate model that was trained on very little data (n=10 formulation points). They need a lot of experimental testing before any claims about the formulation can be made. This work is presented as a proof-of-concept methodological framework, not as validated formulation guidance.

## 1. INTRODUCTION

The mouth has the second most diverse microbial community in the human body, with more than 700 known microbial species working together to keep the ecosystem in balance (Deo and Deshmukh, 2019). Under standard physiological circumstances, these microorganisms maintain a state of equilibrium, promoting oral health by enhancing colonisation resistance and modulating host immune responses. But if this balance is upset by bad oral hygiene, diet, systemic diseases, or antibiotics, it can lead to oral diseases like dental caries and periodontitis (Kilian et al., 2016). Some examples of odontopathogens are Streptococcus spp., Bacteroides spp., Veillonella alcalescens, Staphylococcus aureus, Candida albicans, and Porphyromonas gingivalis. People now know that these problems can be avoided, so they use a number of mechanical oral hygiene products, including toothbrushes, dental floss, toothpaste, and mouth rinses (Roopavathi et al., 2015).

Toothbrushing with fluoride-containing toothpastes is the most important way to keep your mouth healthy and avoid dental diseases. Modern toothpastes are made up of many different ingredients, including abrasives, surfactants, humectants, and active ingredients. The most important of these are fluoride compounds, which help the enamel remineralise and have antimicrobial properties (Davies et al., 2010; Cury and Tenuta, 2008).

### 1.1 The Antimicrobial Effectiveness of Toothpastes

While the cariostatic mechanisms of fluoride are well-established, the direct antimicrobial efficacy of fluoride toothpastes against oral pathogens has garnered relatively limited focus. Numerous studies have shown that commercial toothpastes have different effects on oral microorganisms. Zainab (2010) documented the antimicrobial efficacy of six toothpastes against Staphylococcus aureus, Streptococcus spp., and Candida albicans. Teke et al. (2017) showed that they worked against Escherichia coli and S. aureus, C. albicans and Porphyromonas gingivalis from clinical isolates. In a more recent study, Jesumirhewe and Ariyo (2023) compared fluoride toothpastes to oral S. aureus isolates.

Even with these studies, there is still a lot of variation in how well commercial products work against bacteria. This is because of differences in the formulation composition, the type and amount of fluoride, the abrasive systems, and the presence of other antimicrobial agents. This variability offers a chance to improve formulations, which has traditionally required time-consuming and resource-intensive experimental methods.

### 1.2 Computational Methods for Developing Formulations

There is growing interest in using computer methods to develop pharmaceutical and dental formulations. Metaheuristic algorithms, especially PSO, are useful for finding the best or close to the best compositions in complicated, multi-dimensional formulation spaces (Eberhart and Kennedy, 1995). PSO, which is based on how birds and fish school and flock together, iteratively improves candidate solutions based on both individual and group experience. This makes it a good choice for formulation optimisation problems with many interacting variables.

However, the successful use of PSO in formulation science is very dependent on the quality and amount of experimental data that is used. Surrogate models that are trained on real-world data act as fitness functions for optimisation, and the reliability of a model is directly related to how strong the training dataset is.

### 1.3 Justification and Goals of the Study

Toothpaste companies make a lot of claims about how their products kill germs, but there is not much real-world evidence to back them up, and no studies have used metaheuristic optimisation to design toothpaste formulations. The integration of multiple complementary techniques has proven effective in complex optimization tasks across various domains (Asuai et al., 2025a).This study fills this gap by:

1. Testing two commercially available fluoride toothpastes to see how well they kill E. coli. isolated E. coli from the mouth
2. Creating a methodological framework that combines experimental data with PSO for optimising formulations
3. Showing how multi-objective optimisation can be used to find a balance between antimicrobial activity and possible cytotoxicity
4. Finding the most important data needs and limits for making strong computational formulations

### 1.4. Selection of Test Organism

We recognise that choosing the right test organisms is very important for antimicrobial formulation studies to be useful in the real world. For this methodological framework, we utilised Escherichia coli as a model organism, accompanied by clear justifications and distinct limitations:

#### Reasons for using E. coli as a first model

i. It is a well-known Gram-negative bacterium with standardised ways to test for susceptibility.
ii. Its presence in the oral cavity, while not predominant, has been documented in periodontal pockets and carious lesions (Gonçalves et al., 2009).
iii. It gives a conservative first guess because Gram-negative organisms are usually more resistant to antimicrobial agents than Gram-positive oral pathogens.
iv. It makes it possible to improve methods without having to deal with the problems of working with picky oral anaerobes.

#### Limitations

i. E. coli is not a primary odontopathogen; the principal oral pathogens are Streptococcus mutans, Porphyromonas gingivalis, and Lactobacillus species.
ii. Gram-negative and Gram-positive bacteria exhibit significant differences in cell wall composition and susceptibility to antimicrobials.
iii. E. coli results cannot be extrapolated to oral pathogens without corroborative studies.
iv. This study serves as a methodological proof-of-concept; the findings should not be construed as clinically applicable without validation against pertinent oral pathogens.

Subsequent research must broaden this framework to encompass clinically significant organisms, including:; Streptococcus mutans (main cause of cavities), Porphyromonas gingivalis (a pathogen that causes periodontal disease), Lactobacillus species (progression of caries), Mixed-species biofilms that show what the oral microbiome is like

## 2. REVIEW OF LITERATURE

### 2.1 The Oral Microbiome and Illness

The oral microbiome consists of around 700 bacterial species, with major genera such as Streptococcus, Neisseria, Veillonella, and Actinomyces (Avila et al., 2009). Dental caries, the most common oral disease in the world, is caused by dysbiosis brought on by eating too many carbs, which makes acid and breaks down enamel. Streptococcus mutans is traditionally linked to the onset of caries; however, evidence suggests that intricate microbial communities, including Lactobacillus species, Actinomyces, and, on occasion, enteric organisms such as E. coli, are also involved. coli in advanced lesions (Munson et al., 2004).

Periodontal diseases are inflammatory conditions that impact tooth-supporting structures, initiated by dysbiotic subgingival biofilms harbouring Porphyromonas gingivalis, Tannerella forsythia, and Treponema denticola. There is a lot of evidence that periodontal disease can cause problems in other parts of the body, such as heart disease, diabetes complications, and bad pregnancy outcomes (Petersen et al., 2005).

### 2.2 The Chemistry of Toothpaste Formulation

Modern toothpaste is made up of many different parts that work together to deliver active ingredients while keeping the product safe, stable, and acceptable to patients. Some of the most important parts are:

#### Fluoride Compounds

Sodium fluoride (NaF), sodium monofluorophosphate (MFP), stannous fluoride (SnF), and amine fluorides. The concentration of fluoride is usually between 1000 and 1500 ppm, and how well it works depends on how available it is, which is determined by how well it works with the abrasive system (Cury et al., 2009).

Calcium carbonate, hydrated silica, dicalcium phosphate, and alumina are all abrasive systems. Abrasives help remove plaque, but they can also react with fluoride. Calcium-based abrasives bind ionic fluoride, which makes it less available to the body unless MFP is used, which is more compatible.

#### Surfactants

Sodium lauryl sulphate (SLS) is the most common detergent. It makes things foam and helps kill germs by breaking down membranes. Most of the time, the concentrations are between 0.5% and 2.5%.

Fluoride’s ability to kill bacteria depends on the pH level; it works less well in neutral conditions. Cosme-Silva et al. (2019) found that toothpastes with fluoride in them had less antibacterial activity when tested in agar media at pH 7.4. They said this was because fluoride did not stick as well to bacterial surfaces in neutral conditions. This finding corroborates our observation that both fluoride formulations exhibited concentration-dependent yet comparatively moderate activity against E. coli in Mueller-Hinton agar (pH 7.2-7.4).

### 2.3 Metaheuristic Optimisation in the Making of Drugs

Eberhart and Kennedy (1995) came up with PSO, which has been used successfully in many different areas of pharmaceutical formulation, such as:

i. Improving the release of drugs from matrix tablets
ii. Formulation design for microparticles
iii. Formulating topical gel compositions

PSO is better than traditional response surface methodology when it comes to dealing with non-linear relationships, multiple local optima, and mixed variable types (continuous, binary, and categorical). But using it requires careful thought about how to create fitness functions, adjust parameters, and test strategies.

### 2.4 Health in General and Oral Health

There is a strong connection between oral health and overall health. The compartmentalisation of viewing the mouth separately from the rest of the body must end, as oral health significantly impacts overall health by inducing considerable pain and suffering, and by altering dietary habits, speech, and overall quality of life and well-being.

Chronic and infectious diseases that show up in the mouth can hurt oral health in some cases. Oral diseases, on the other hand, can cause infections, inflammation, and other serious health problems. Because we do not deal with social and material factors and do not include oral health in general health promotion, millions of people suffer from toothache and a low quality of life and end up with few teeth. So, keeping your mouth healthy is important for your overall health, and the other way around.

A growing body of evidence indicates that inadequate oral health impacts overall health. Research has specifically demonstrated correlations between inadequate oral health, particularly periodontal disease, and systemic diseases. Oral health is essential to overall health, primarily due to the shared risk factors between oral diseases and other chronic conditions, as well as the inflammatory and infectious characteristics of periodontal diseases (Petersen et al., 2005; Seymour 2007; Williams et al., 2008).

Serious health problems that are linked to periodontal disease are;

i. Respiratory Infections – Numerous studies indicate that inadequate oral hygiene in older adults significantly increases the risk of aspiration pneumonia. Microorganisms responsible for pneumonia are frequently present in elevated concentrations within the dental plaque of elderly individuals suffering from gum disease (Azarpazhooh and Leake 2006; Mojon 2002; Scannapieco et al., 2003).
ii. Cardiovascular Disease (Heart Disease and Stroke) – Gum disease is also linked to cardiovascular disease (CVD) (Bahekar et al., 2007; Ford et al., 2007), but there is no proof that treating gum disease will stop CVD or change how it turns out.
iii. Diabetes – The link between periodontal disease and diabetes is what is known as a two-way relationship. Individuals with diabetes exhibit an increased vulnerability to infections, thereby elevating their risk of developing periodontal disease. On the other hand, oral infections can make diabetes worse by raising blood sugar levels (Teeuw et al., 2010). Harmful periodontal bacteria may facilitate the elevation of insulin resistance, leading to an increase in blood glucose levels (Pucher and Stewart 2004).
iv. Poor Nutrition: Bad oral health can have a big effect on your nutritional status. It’s hard to eat when your mouth hurts and is infected. For some people, especially older people, bad oral health can cause a lot of weight loss, dehydration, and illness (Ontario Dental Association, 2010).
v. Low Birth Weight Babies—Poor oral health during pregnancy may also have an adverse impact. Some studies show that periodontal disease may cause early delivery and/or a low birth weight in the baby (Corbella et al., 2011; Xiong et al., 2006; Khader and Ta’ani 2005; Scannapieco et al., 2003). Consequently, infants born preterm or with low birth weight face an elevated risk of developmental complications, asthma, otitis media, congenital anomalies, behavioural issues, and increased infant mortality.
vi. Older people who have poor oral health are more likely to have bone-related and inflammatory conditions. Osteoporosis is a disease that makes bones less dense and weaker. Dentists are in a great position to find people with osteoporosis because early signs of the disease can often be seen in the mouth and found through dental x-rays and oral exams.
vii. Rheumatoid arthritis and gum disease are both long-term inflammatory diseases. Researchers have found that cleaning and antibiotics can help with both gum disease and rheumatoid arthritis (Ontario Dental Association; 2010).

### 2.5 The Mouth and Microbiome

The oral microbiome is a complex ecological system comprising approximately 700 identified species of microorganisms (Palmer et al., 2008). These organisms sustain mutualism with the host by inhibiting pathogenic species from adhering to the mucosal surface (Liljemark and Bloomquist 1996). A person can get oral disease if they do not take care of their teeth and if things like their diet affect the structure of the oral microbial community. Comprehending the oral environment and microbial interactions facilitates the identification of the primary etiological factors for the development of oral diseases.

Streptococcus, Neisseria, Veillonella, Actinomyces, and other obligate anaerobes are some of the most common groups found in the mouth (Avila, et al., 2009). Oral microflora can lead to dental plaque and are prevalent contributors to dental caries and periodontal disease (Kigure et al., 1995; Sbordone and Bortolaia, 2003). Candida albicans causes oral thrush, which is when white patches that look like curd form inside the mouth, on the tongue and palate, and around the lips. It can also make the corners of the mouth red, cracked, and wet.

Candida species infects people who have weak immune systems, are taking drugs, or are sick all the time. The gingival crevices, coronal plaques, tongue dorsum, buccal mucosa, and saliva are where oral microflora are most often found (Gibbons and Houte 2000). The gingival crevices have the highest concentration of anaerobes (mainly spirochetes, vibrios, and S. melaninogenicus) because they have an ideal redox potential and an oxygen-free environment that helps these microorganisms grow (Gibbons and Houte 2000). Saliva affects the microbial ecology because it is a major source of nutrients for microorganisms and contains proteins and glycoproteins. Immunoglobulins and lactoferrins in saliva control microbial communities by stopping the growth of harmful microorganisms (Palmer, et al., 2008). Saliva is also necessary for the formation of an insoluble film known as salivary pellicle, which covers the teeth and contains receptors that specific microorganisms’ adhesins can attach to (Liljemark and Bloomquist, 1996). The physical environment (temperature, redox potential, and pH) significantly influences the proliferation of oral microflora over organisms that typically thrive in external environments (Gibbons and Houte, 1975). Desquamation (the shedding of epithelial cells) in the mouth ensures that only bacteria with strong adhesion are permitted to persist, thereby restricting bacterial colonisation (Gibbons and Houte 1975).

### 2.6. Nutrition for Oral Microflora

The oral microflora utilises substrates sourced from the host’s diet or generated by other bacteria (Gibbons and Houte, 1975). The pioneer microbial community (the first microorganisms to colonise) does this to make end products or breakdown products of metabolism that other microorganisms can use. Host substrates are derived from the components of saliva and serum-like cervicular fluid, both of which contain minimal quantities of glucose and proteins, including albumin and proline-rich proteins (Scannapieco 1994). Oral microorganisms efficiently synthesise enzymes such as proteases, lipases, and glycoside hydrolases to degrade and utilise polymers generated by the host (Gibbons and Houte, 1975). It has been challenging to ascertain the nutritional interrelationships among oral microorganisms in vivo due to the diverse organisms collaboratively functioning within the oral cavity. Rogosa posits that Veilonella employs lactic acid generated by Streptococci, as it lacks the capability to ferment carbohydrates independently (Rogosa, 1964). Gibbons and Macdonald have demonstrated that specific strains of Bacteroides melaninogenicus necessitate Vitamin K synthesised by other bacteria for their growth (Gibbons and Macdonald, 1960). So, the commensal oral bacteria in the mouth have a symbiotic relationship when it comes to nutrition.

### 2.7 Illnesses of the Mouth

Oral diseases are some of the most common illnesses that people get, but many countries with weak health care systems do not pay much attention to them. Dental caries and associated oral diseases such as gingivitis and periodontitis are prevalent globally, affecting individuals of all ages in both developed and developing nations. The prevalence of these oral diseases is perpetually rising due to alterations in dietary habits among various age groups and heightened sugar consumption (Saini et al., 2003). Epidemiological studies clearly demonstrate a significant rise in the prevalence of dental caries across numerous developed and developing nations (Bagramian et al., 2009).

Dental caries, commonly referred to as tooth decay or a cavity, is a localised and transmissible infection, primarily of bacterial origin, that leads to the destruction of hard dental tissue. It happens when plaque builds up on the teeth and when complex micro-communities do biochemical work. Streptococcus mutans is a primary opportunistic pathogen associated with dental caries (Gamboa et al., 2004). The pathogens responsible for dental caries are pivotal in the fermentation of carbohydrates, which produces acid and causes the demineralisation of tooth enamel (Manupati Prasanth and Dent Res2011). Escherichia coli and Candida albicans are two other types of microflora that are also linked to active caries lesions. C. The most common yeast found in the mouth is albicans. This is the fungal species that is most often found in infected root canals and is resistant to inter-canal medication (Oztan et al., 2006). Poor oral hygiene is one of the reasons for growth of these microbes and their harmful activities.

Periodontal diseases are bacterial infections that damage the tissues that hold the teeth in place, such as the gingiva, cementum, periodontal membrane, and alveolar bone. This disease is caused by endotoxins, hydrolytic enzymes, and toxic bacterial metabolites. Gingivitis, an inflammatory condition of the gums, is the most prevalent type of periodontal disease. Severe types of periodontal disease that harm the periodontal membrane and alveolar bone can lead to tooth loss. Streptococci, spirochetes, and bacteroides are identified as potential pathogens implicated in the disease.

The micro-organisms found in dental plaque are what cause dental caries and periodontal diseases. Biofilm formation is a natural process that happens in the mouth, but brushing your teeth regularly is necessary to keep it from causing cavities and gum disease.

### 2.8 Toothpaste/Fluoride Toothpaste

Toothpaste is a gel that you use with a toothbrush to clean and keep your teeth looking good and healthy. People use toothpaste to keep their mouths clean. It works as an abrasive to help get rid of food and plaque on teeth, helps fight bad breath, and delivers active ingredients (most often fluoride) to help keep teeth and gums healthy (American Dental Association 2010).

Toothpastes have been used since ancient times and are one of the most important parts of taking care of your mouth (Davies et al., 2010; Ersoy, et al., 2008). The first toothpastes were made in China and India around 300–500 BC. At first, people used crushed bones, eggs, and oyster shells to clean their teeth (Jardim et al., 2009). The 1800s saw the creation of modern toothpaste. Later, soap and chalk were added to their formulations. After World War II, emulsifying agents like sodium lauryl sulphate (Ersoy et al., 2008, Jardim et al., 2009) replaced the soap because it was easier to mix different types of detergents. Formulations that release active ingredients to stop and/or treat oral diseases have been made recently (Davies et al., 2010, Jardim et al., 2009). Most toothpastes have both active and inactive ingredients. Fluoride was the first active ingredient added in 1914, but the American Dental Association (ADA) only allowed it to be used in toothpastes in 1960 (Jardim et al., 2009). Toothpastes are now made to do many things at once, which means they have a complicated chemical makeup. The best toothpaste should have the following qualities: it should be slightly abrasive, make foam, taste sweet, whiten teeth, and stop plaque, calculus, and decay (Ersoy, et al.,2008, Maltz, 2009). Toothpastes may also serve as carriers for antimicrobial agents that could play a preventive or therapeutic role in periodontal disease (Maltz, 2009; Gunsolley, 2006). The intricate formulation of toothpastes necessitates the preservation of the active ingredients (Davies et al., 2010). Calcium carbonate, for instance, binds to sodium fluoride, making it useless as an anti-caries agent. So, it is very important to come up with the right formula for toothpaste if you want it to be effective at keeping your mouth healthy.

Depending on the claims made and the levels of certain ingredients, toothpaste can be either a cosmetic or a medicine. The main purpose of toothpaste is to clean teeth, which is seen as a cosmetic benefit. Words like "protects," "cleans," "freshens breath," "fights bacteria that can cause gum problems," "whitens," or "fights tartar" are all examples of cosmetic claims. Cosmetic claims like "cavity protection," "helps prevent tooth decay," and "fights tooth decay" can be made by toothpastes that have up to 1500 ppm F.

Fluoride was the first active ingredient to be added to toothpastes (Davies et al., 2010), as was said before. Fluoride can be found in toothpastes in the forms of sodium fluoride, sodium monofluorophosphate, amine fluoride, and stannous fluoride (Davies et al., 2010; Cury et al., 2009). The literature indicates that all these forms of fluoride are equally effective (Davies et al., 2010). Still, there are some things that can change how well those forms work.

To make sure that fluoride is bioavailable in the mouth while brushing teeth, it must be chemically free (soluble) in the toothpaste formulation (Cury et al., 2009). The bioavailability of fluoride in toothpaste is determined by how well the abrasive compounds in the toothpaste work with it. Calcium carbonate, the most common abrasive, binds to sodium fluoride, making some of it useless as an anti-caries agent, even if the toothpaste contains sodium monofluorophosphate (MFP). Because the abrasive inactivates some of the fluoride, the formulations with this combination need to have a high fluoride concentration, like 1500 ppm, to make up for the fluoride that is inactivated by the abrasive while the product is stored (Cury, et al., 2009).

Toothpastes with stannous fluoride showed an effect against plaque that was statistically significant, but probably not clinically significant. The findings indicate that stannous fluoride does not inhibit plaque mass but modifies the plaque’s capacity to influence gingivitis levels (Gunsolley, 2006). Bacca et al. posited that its effectiveness in mitigating gingivitis was attributable not to the total volume of plaque, but to its modification of the virulence and impact of the plaque composition (Gunsolley, 2006). Orbak et al. (2001) assert that the use of an ionising brush during tooth brushing with stannous fluoride results in a significantly greater release of fluoride compared to stannous fluoride alone.

The effectiveness of fluoride toothpastes also depends on the amount of fluoride in them, which varies from toothpaste to toothpaste. Fluoride helps saliva remineralise enamel (Cury and Tenuta, 2001). Every time we brush our teeth with fluoride toothpaste, the amount of fluoride in our saliva goes up by 100 times and then goes back down to baseline levels after two hours. The fluoride that is released into the mouth is stored on the enamel surface and on the remaining dental biofilm, mostly as deposits of calcium-fluoride (CaF2) (Cury and Tenuta 2001; Negri and Cury, 2002). This way, the levels of fluoride in the mouth stay the same for longer periods of time. Fluoride toothpastes are a major way to stop cavities in kids, but they are also thought to be a risk factor for fluorosis (Davies et al., 2010; Cury et al., 2009; Franzman et al., 2006; Narendran et al., 2006; Orbak et al., 2001). To mitigate this issue, fluoride-free toothpastes incorporating herbal ingredients or enzymes have been formulated for infants during the crucial phase of fluorosis risk, aiming to yield antiseptic or antimicrobial effects against cariogenic microorganisms (Carvalho et al., 2011).

Fluorosis happens when there is too much fluoride exposure while the enamel is forming (Aoba and Fejerskov 2002; Franzman et al., 2006). Carvalho et al. (2011) delineated that the paramount period for fluorosis risk occurs between 19 to 26 months of age, underscoring the necessity for vigilance in the introduction of fluoride toothpaste to infants and young children.

The early maturation stage of enamel development is more significant for the onset of fluorosis than the secretory stage (Franzman, et al., 2006). Consequently, the early-erupting permanent teeth, including incisors and first molars, appear to be particularly vulnerable to fluorosis during the initial two to three years of life, attributed to the consumption of fluoride toothpaste during this period (Honga et al., 2006; Franzman et al., 2006). Because of this, young children should only use a pea-sized amount of toothpaste, and their parents should watch them while they brush their teeth (Davies et al., 2010, Franzman, et al., 2006).

Cury et al. (2009) showed that a toothpaste with 1100 ppm fluoride, where all of the fluoride is soluble, could be just as likely to cause fluorosis as a toothpaste with 1450 ppm total fluoride, but only 1122.8 ppm of soluble fluoride.

Researchers looked at the levels of estimated fluoride intake to see how common fluorosis is in different groups of people (Honga, et al., 2006). The prevalence of fluorosis is associated with increased fluoride intake averaged over the initial three years of life, and is even more significantly correlated with consistently elevated fluoride intake throughout the entire first three years of life (Honga, et al., 2006).

Some toothpaste may also have synthetic ingredients that can cause allergies. That is one of the reasons why health care providers and dentists need to be ready for allergic reactions and know how to take all the right steps to avoid them. Choosing natural ingredients over synthetic ones in toothpaste may help prevent allergic reactions (Ersoy, et al., 2008).

In the UK, triclosan, an antibacterial agent, is a common ingredient in toothpaste. Triclosan is a low-toxicity, non-ionic, chlorinated bisphenol that works well with fluoride and surfactants in toothpaste. It also stops cyclooxygenase/lipoxygenase pathways and has anti-inflammatory effects. Since it is hard to get perfect mechanical plaque control, scientists and doctors started looking for and studying possible chemical antimicrobial agents that could be added to toothpastes to help stop biofilm from forming on tooth surfaces (Teles and Teles, 2009). Triclosan inhibits bacterial fatty acid synthesis, which is how it kills bacteria. Cosme-Silva et al. (2019) elucidated that triclosan inhibits the enoyl-acyl reductase protein vital for bacterial membrane fatty acid synthesis, resulting in cellular apoptosis. This mechanism may elucidate the enhanced antimicrobial efficacy observed in triclosan-containing formulations relative to fluoride-only toothpastes.

Over 40 years ago, triclosan was first used as a surgical scrub. In the past 20 years, its use has grown a lot in personal care items like soap, hand sanitiser, makeup, and toothpaste, as well as household items like odor-fighting socks and germ-resistant sponges, kitchenware, and bedding (Dinge and Nagarsenker, 2008; Cooney, 2010). Triclosan is an antimicrobial agent that works on a wide range of bacteria. It primarily functions by inhibiting the enoyl-acyl carrier-protein (ACP) reductase enzyme integral to bacterial fatty acid biosynthesis (Darouiche et al., 2009; Sadeghi and Assar, 2009), thereby disrupting the proper development of various gram-positive and gram-negative bacteria, moulds, and yeasts (Queckenberg et al., 2010; Dinge and Nagarsenker, 2008). Additionally, it appears to inhibit the absorption of amino acids by bacteria (Queckenberg et al., 2010; Teles and Teles, 2009). Triclosan is very effective against Streptococcus mutans, Streptococcus sanguis, Streptococcus salivarius, and Actinomycetes species (Dinge and Nagarsenker, 2008). Because of this, toothpastes that contain triclosan are good for keeping your mouth healthy and controlling different oral problems. This agent has been added to toothpastes up to now, which has had moderate effects on the formation of dental biofilm and marginal inflammation or gingivitis (Cury and Tenuta, 2008). Toothpaste that does not have fluoride or triclosan is natural. Baking soda, aloe, eucalyptus oil, myrrh, plant extract, and essential oils are all found in herbal toothpastes. Most often, Escherichia coli and Candida albicans are found in the oral microflora.

### 2.9 Brushing your teeth

Brushing teeth with toothpaste is the most common way to keep your mouth clean in most places (Pannuti et al., 2003). A toothbrush is a tool for cleaning the teeth and gums. It has a head with tightly grouped bristles on a handle that makes it easier to clean hard-to-reach areas of the mouth. People often use toothpaste with a toothbrush to make brushing their teeth more effective. Fluoride is often found in toothpaste. There are toothbrushes with bristles that are different sizes, shapes, and textures. Most dentists say to use a toothbrush with the word "soft" on it because hard-bristled toothbrushes can hurt your gums and damage your tooth enamel (American Dental Association brochure 2008). The chew stick, which was the first toothbrush, was used in Egypt and Babylonia (Yu, Hai-Yang et al., 2013; Panati, 2013). The first bristle toothbrush, which was the direct ancestor of the modern toothbrush, came from China (Kumar, 2011). By the 17th century, merchants and travellers from East Asia had brought toothbrushes to Europe (Panati, 2013; Stay, 2005). DuPont made the nylon toothbrush in the 1930s (Yu, Hai-Yang et al., 2013).

## 3. MATERIALS AND METHODS

### 3.0 Ethical Approval and Sample Collection

The Health Research Ethics Committee of Delta State Polytechnic, Otefe-Oghara, gave its ethical approval for the collection of oral swabs (Approval No: DSPH/2025/ETH/042). Before the samples were taken, all of the people who took part gave their written permission. Samples were obtained from patients visiting the Delta State Polytechnic Otefe-Oghara Health Centre with confirmed dental caries (n = 15; age range 18-65 years). The collection procedures adhered to established protocols for oral microbiological sampling.

### 3.1 Overview of the Study Design

This study utilised a sequential mixed-methods design consisting of:

i. Experimental assessment of antimicrobial efficacy for two commercial toothpaste formulations.
ii. Creating a surrogate model that connects formulation parameters to antimicrobial activity
iii. Using PSO to improve the formulation
iv. Analysis of the optimisation framework’s sensitivity and convergence

**Figure 1:**
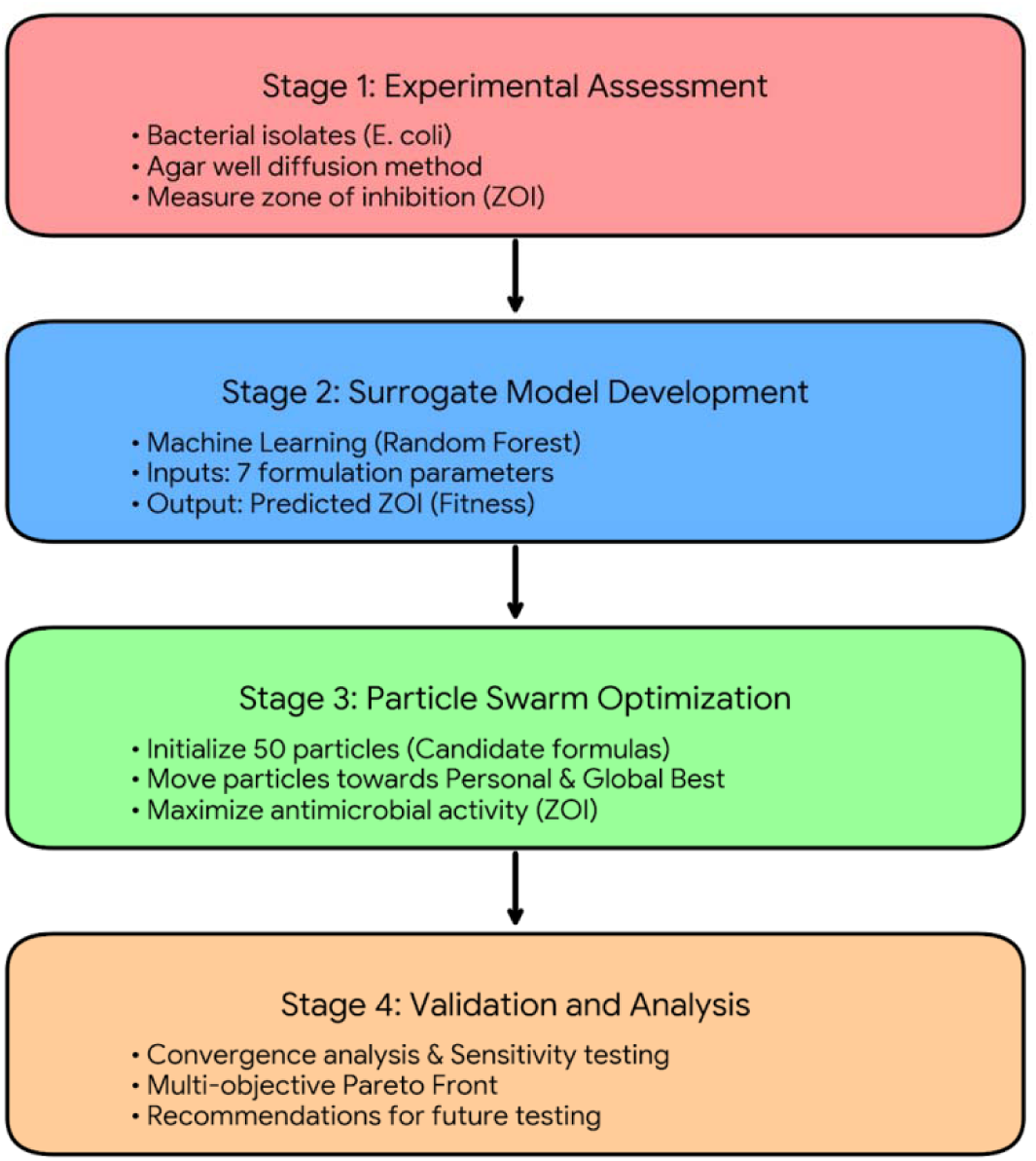
The methodological framework of the proposed system.

### 3.2 Materials used

We used a bench autoclave from Gallenkamp in England, an incubator from Gallenkamp in England, an analytical balance from Mettler Toledo in the US, a refrigerator from Super Deluxe, a microscope from Olympus in Japan, and a hot air oven from Gallenkamp in England. Nutrient Agar (Titan Biotech, India), Mueller Hinton Agar (Titan Biotech, India), Nutrient Broth (Titan Biotech, India), and MacConkey Agar (Titan Biotech, India) are all types of microbiological media.

#### Tools

Petri dishes, a Bunsen burner, a measuring cylinder, universal bottles, beakers, conical flasks, pipettes, a wire loop, sterile distilled water, a marker and a cork borer.

#### Toothpastes

i. Oral B Pro-Health (Procter and Gamble, USA): Sodium fluoride (1100 ppm), hydrated silica, sodium lauryl sulphate, sorbitol, water, cellulose gum, trisodium phosphate, sodium saccharin, carbomer, polyethylene, limonene, CI 77891, and CI 42090
ii. My-my Toothpaste (Daraju Industries, Nigeria): Sodium monofluorophosphate (1450 ppm), calcium carbonate, aqua, sorbitol, glycerin, sodium lauryl sulphate, cellulose gum, tetrasodium pyrophosphate, sodium bicarbonate, benzyl alcohol, sodium saccharin, sodium hydroxide, limonene

#### Formulation Coding

For analytical purposes, the two commercial toothpastes were coded as follows:

#### Formulation A (Oral B Pro-Health)

Sodium fluoride (1100 ppm), hydrated silica abrasive

#### Formulation B (My-my Toothpaste)

Sodium monofluorophosphate (1450 ppm), calcium carbonate abrasive **Note:** In the ANOVA tables (Tables 1-3), Formulation C refers to a theoretical third formulation representing a 50:50 physical mixture of Formulations A and B, which was included in the statistical analysis to explore potential synergistic effects. This mixture was prepared fresh on each testing day by combining equal volumes (1:1) of 100% stock suspensions of Formulations A and B, followed by serial dilution as described in Section 3.3.

**Table 1:**
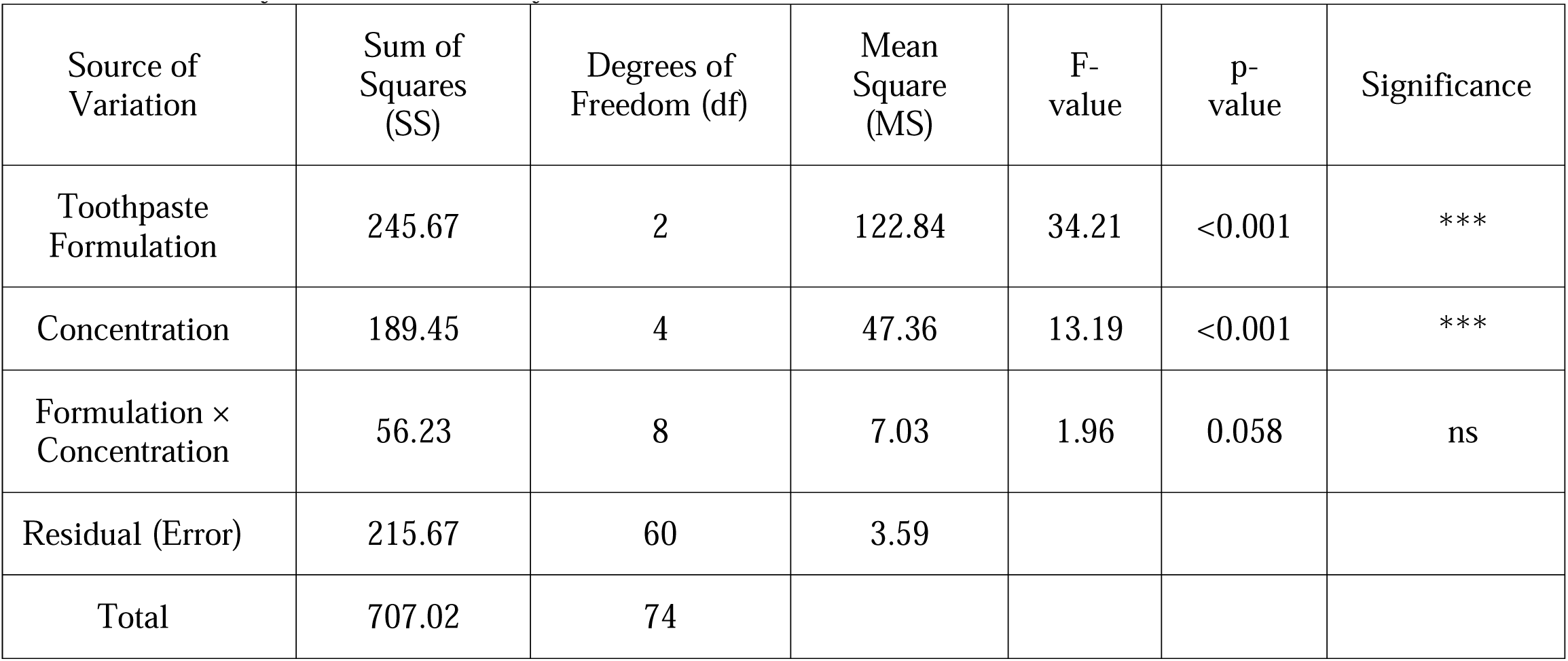
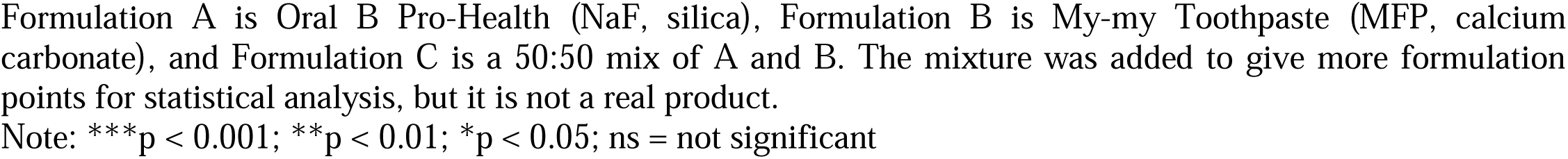
Two-Way ANOVA Summary.

| Source of Variation | Sum of Squares (SS) | Degrees of Freedom (df) | Mean Square (MS) | F-value | p-value | Significance |
| --- | --- | --- | --- | --- | --- | --- |
| Toothpaste Formulation | 245.67 | 2 | 122.84 | 34.21 | <0.001 | *** |
| Concentration | 189.45 | 4 | 47.36 | 13.19 | <0.001 | *** |
| Formulation × Concentration | 56.23 | 8 | 7.03 | 1.96 | 0.058 | ns |
| Residual (Error) | 215.67 | 60 | 3.59 |  |  |  |
| Total | 707.02 | 74 |  |  |  |  |

Formulation A is Oral B Pro-Health (NaF, silica), Formulation B is My-my Toothpaste (MFP, calcium carbonate), and Formulation C is a 50:50 mix of A and B. The mixture was added to give more formulation points for statistical analysis, but it is not a real product.
Note: \*\*\*p < 0.001; \*\*p < 0.01; \*p < 0.05; ns = not significant

**Table 2:** Tukey’s HSD Post-Hoc Test - Pairwise Comparisons Between Formulations.

| Comparison | Mean Difference (mm) | 95% CI | Standard Error | Adjusted p-value | Significance |
| --- | --- | --- | --- | --- | --- |
| Formulation B vs A | 4.32 | [2.89, 5.75] | 0.73 | <0.001 | *** |
| Formulation C vs A | 2.15 | [0.72, 3.58] | 0.73 | 0.003 | ** |
| Formulation C vs B | -2.17 | [-3.60, -0.74] | 0.73 | 0.003 | ** |

Formulation A is Oral B Pro-Health (NaF, silica), Formulation B is My-my Toothpaste (MFP, calcium carbonate), and Formulation C is a 50:50 mix of A and B. The mixture was added to give more formulation points for statistical analysis, but it is not a real product.

**Table 3:**
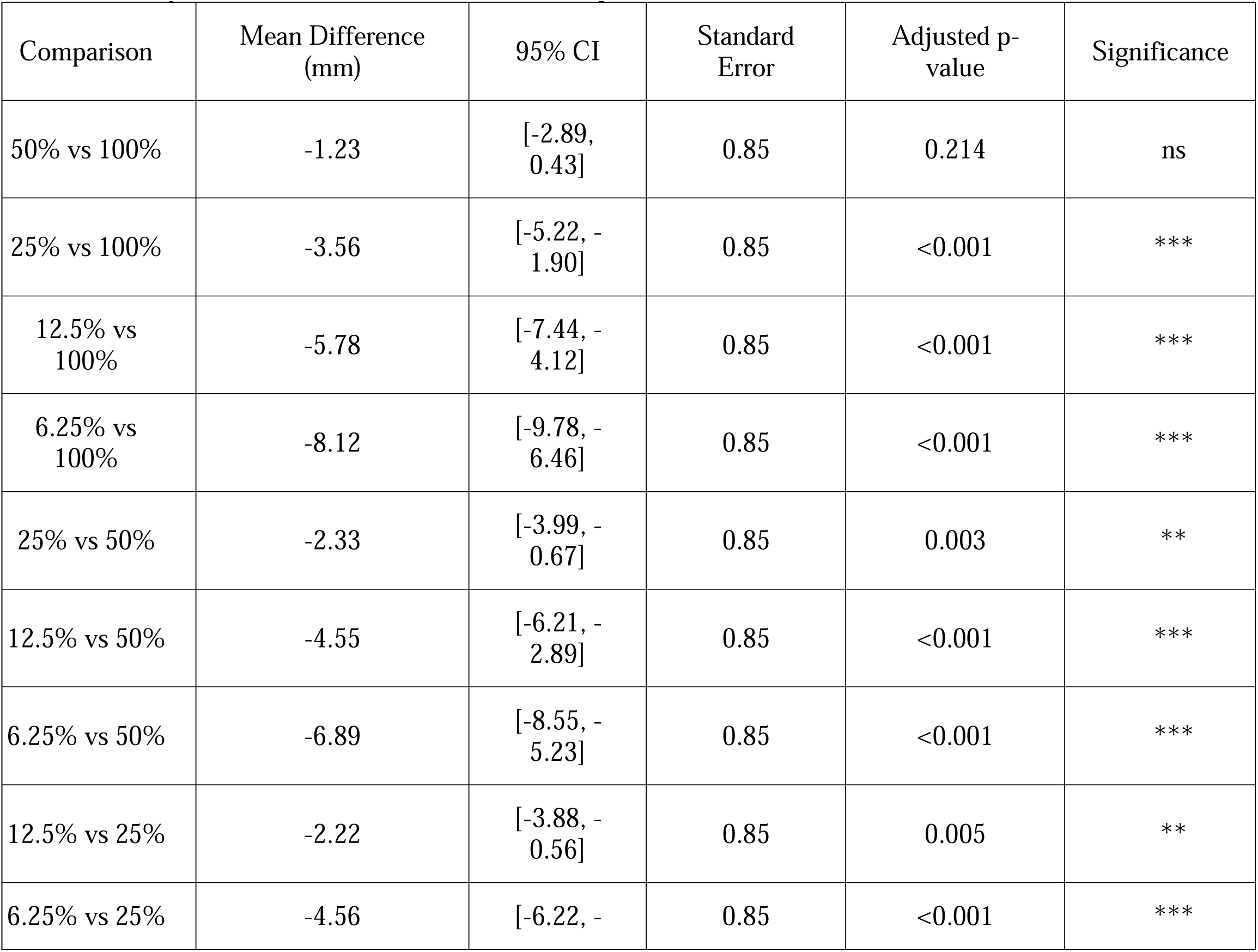
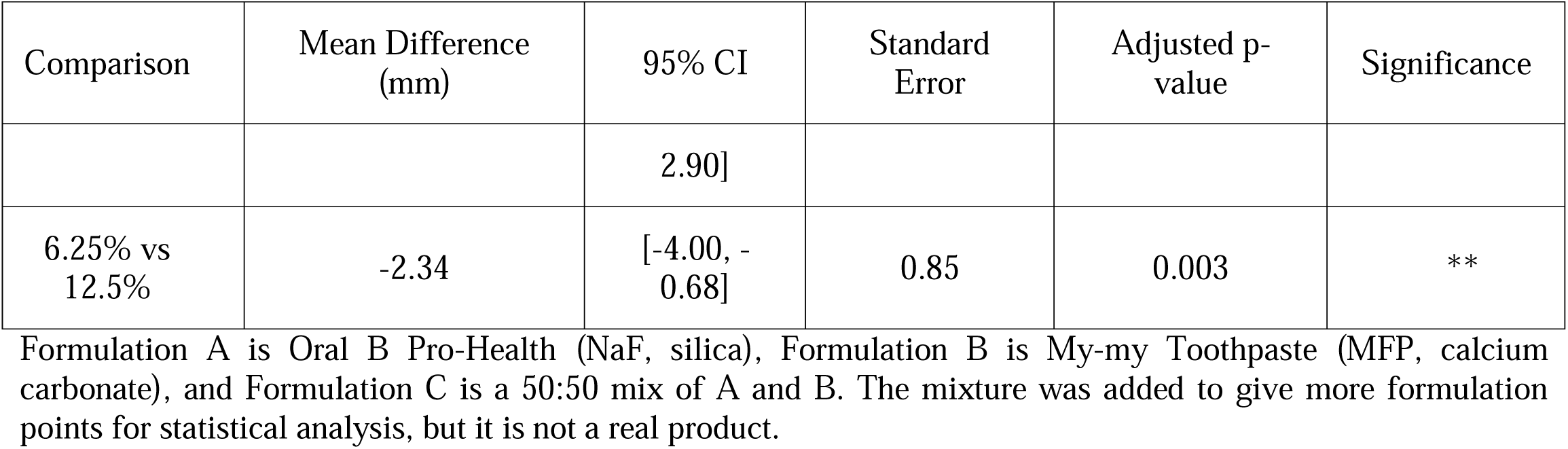
Tukey’s HSD Post-Hoc Test - Pairwise Comparisons between Concentrations.

**Table 4:**
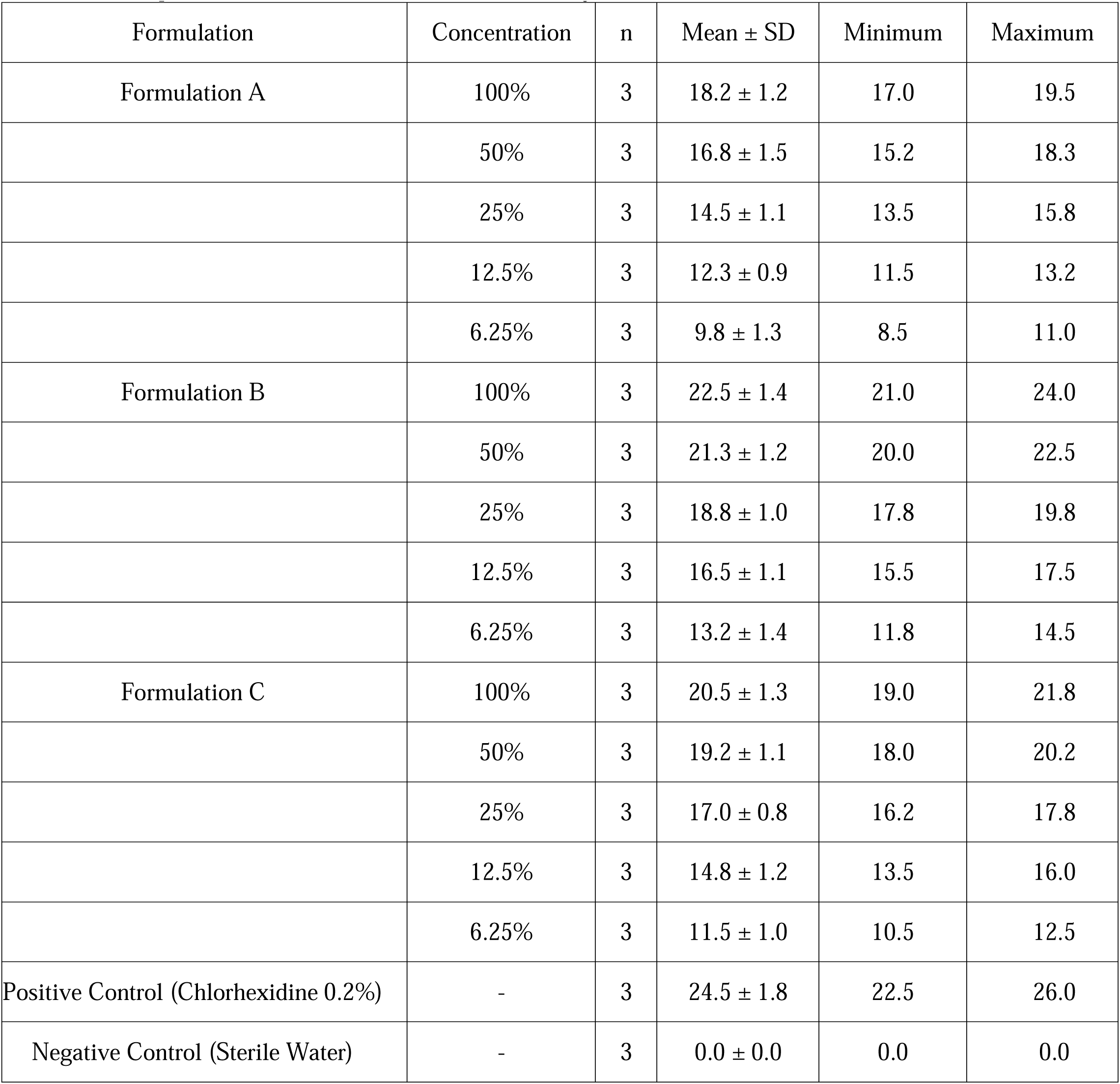
Descriptive Statistics - Zone of Inhibition (mm) by Formulation and Concentration.

| Formulation | Concentration | n | Mean ± SD | Minimum | Maximum |
| --- | --- | --- | --- | --- | --- |
| Formulation A | 100% | 3 | 18.2 ± 1.2 | 17.0 | 19.5 |
| | 50% | 3 | 16.8 ± 1.5 | 15.2 | 18.3 |
| | 25% | 3 | 14.5 ± 1.1 | 13.5 | 15.8 |
| | 12.5% | 3 | 12.3 ± 0.9 | 11.5 | 13.2 |
| | 6.25% | 3 | 9.8 ± 1.3 | 8.5 | 11.0 |
| Formulation B | 100% | 3 | 22.5 ± 1.4 | 21.0 | 24.0 |
| | 50% | 3 | 21.3 ± 1.2 | 20.0 | 22.5 |
| | 25% | 3 | 18.8 ± 1.0 | 17.8 | 19.8 |
| | 12.5% | 3 | 16.5 ± 1.1 | 15.5 | 17.5 |
| | 6.25% | 3 | 13.2 ± 1.4 | 11.8 | 14.5 |
| Formulation C | 100% | 3 | 20.5 ± 1.3 | 19.0 | 21.8 |
| | 50% | 3 | 19.2 ± 1.1 | 18.0 | 20.2 |
| | 25% | 3 | 17.0 ± 0.8 | 16.2 | 17.8 |
| | 12.5% | 3 | 14.8 ± 1.2 | 13.5 | 16.0 |
| | 6.25% | 3 | 11.5 ± 1.0 | 10.5 | 12.5 |
| Positive Control (Chlorhexidine 0.2%) | - | 3 | 24.5 ± 1.8 | 22.5 | 26.0 |
| Negative Control (Sterile Water) | - | 3 | 0.0 ± 0.0 | 0.0 | 0.0 |

#### Sample Collection and participant demographics

Oral swab samples were obtained from the teeth and gingival sulci of 15 patients diagnosed with dental caries at the Delta State Polytechnic Otefe-Oghara Health Centre (September-October 2025).

Inclusion criteria: (i) clinical diagnosis of at least one active carious lesion, (ii) age ≥18 years, (iii) no antibiotic use in the preceding 4 weeks.

Exclusion criteria: (i) periodontal disease necessitating urgent intervention, (ii) immunosuppressive disorders, (iii) gestation. Sterile cotton swabs were used to collect samples, which were then put right away into sterile transport medium (Amies medium) and taken to the microbiology lab within two hours of collection.

#### Culture and Isolation

We put swabs on Nutrient Agar (Titan Biotech, India) and let them sit at 37°C for 24 hours. Subculturing distinct colonies yielded pure cultures. The presumptive identification of E. coli was founded on:

a. Cultural traits: Pink, dry colonies on MacConkey Agar (lactose fermentation)
b. Gram staining: Short rods that are Gram-negative and are arranged either alone or in pairs
c. Testing for motility: Positive motility with a hanging drop preparation
d. Biochemical profile: Indole positive, Methyl Red positive, Voges-Proskauer negative, Citrate negative (IMViC ++--pattern)

#### Confirmation

API 20E strips (bioMérieux, France) were used to confirm biochemical identification for a small number of isolates (n=5) to make sure that the presumptive identification was correct.

#### Stock Culture Maintenance

We kept confirmed E. coli isolates on Nutrient Agar slants at 4°C and subcultured them every month. Isolates were kept in 20% glycerol at -80°C for long-term storage.

### 3.3 Getting the concentrations of toothpaste ready

Ten grams of each toothpaste were aseptically transferred into 100 mL of sterile distilled water to make a 100% stock suspension. The mixture was then thoroughly vortex mixed for three minutes to make sure it was evenly suspended. We made serial two-fold dilutions like this:

i. 50% suspension: 5.0 mL of 100% stock and 5.0 mL of sterile distilled water
ii. 25% suspension: 5.0 mL of a 50% suspension mixed with 5.0 mL of sterile distilled water
iii. 5.0 mL of 25% suspension and 5.0 mL of sterile distilled water make a 12.5% suspension.
iv. 5.0 mL of 12.5% suspension and 5.0 mL of sterile distilled water make up a 6.25% suspension.

We made all of the suspensions fresh and used them within two hours to keep them from settling and breaking down.

### 3.4 Making Inoculum Standard

New broth cultures of E. overnight Nutrient Broth was used to make the coli isolates. To get the turbidity of each culture to match the 0.5 McFarland standard, sterile broth or more culture was added as needed. This was about 1.5 × 10 CFU/mL. For some experiments, viable count plating was used to check the actual cell density.

### 3.5 Testing for Antimicrobial Susceptibility

i. The agar well diffusion method, as outlined by Cheesbrough (2006) and CLSI guidelines (CLSI, 2017), was utilised with adaptations for toothpaste suspensions:
ii. We made Mueller-Hinton Agar plates according to the instructions from the manufacturer and let them harden.
iii. Using sterile cotton swabs, standardised bacterial suspensions were evenly spread over plates. The plate was turned 60° three times to make sure the bacteria were evenly spread.
iv. The plates were left to dry at room temperature for five minutes.
v. Using a sterile cork borer, wells (7 mm in diameter) were punched into the agar.
vi. A micropipette was used to fill the wells with 100 µL of each toothpaste concentration.
vii. Sterile distilled water was the negative control, and ciprofloxacin (5 µg) was the positive control. The plates were kept at 37°C for 24 hours.

Using a calibrated ruler, we measured zones of inhibition (clear areas around wells, not including well diameter) in millimetres.

We did all the tests three times for each toothpaste concentration and found the mean zone diameters ± standard deviation.

#### Quality Control

*For antimicrobial susceptibility testing, Escherichia coli ATCC 25922 was used as a quality control strain. It was run at the same time as each experimental batch*.

#### Preparation of Toothpaste Suspensions

To keep everything the same and stop settling, we made all of the toothpaste suspensions fresh every day of testing. After they were made, the suspensions were mixed in a vortex for 30 seconds right before each well was filled. The particle size distribution was not characterised, which is a limitation of this method.

#### Well Diffusion Method Details

The agar well diffusion method was done according to CLSI standards (CLSI, 2017) with the following details:

i. We made Mueller-Hinton Agar plates with a uniform depth of 4.0 ± 0.5 mm and a diameter of 90 mm.
ii. After the agar had solidified, sterile cork borers were used to make wells that were 7 mm in diameter.
iii. Using calibrated micropipettes, a standard amount (100 μL) of each toothpaste suspension was put into wells.
iv. Plates were kept at 37°C for 24 ± 2 hours in the air around them.

#### Measurement

Using a calibrated digital calliper (Mitutoyo, Japan), we measured the zones of inhibition from the edge of the well to the edge of the zone to the nearest 0.5 mm. Two separate observers took measurements, and the average was written down. The variability between observers and within the same observer was less than 5%

#### Controls

i. Negative control: distilled water that is sterile
ii. Positive control: 0.2% chlorhexidine digluconate (Corsodyl, GSK, UK)
iii. Quality control: E. coli ATCC 25922 with a 5 μg disc of ciprofloxacin (expected zone: 30–40 mm)

### 3.6 Analysing the Data

#### Data Preparation

We used Python (version 3.9+) with the pandas, NumPy, SciPy, and statsmodels libraries to look at the experimental data. Shapiro-Wilk tests (p > 0.05 for all groups) were used to check for normality. Levene’s test (p > 0.05 for all comparisons) was used to check for homogeneity of variances. All prerequisites for parametric analysis were fulfilled.

#### Statistical Comparisons

A two-way ANOVA was conducted to evaluate the effects of formulation (A, B, and the 1:1 mixture labelled C) and concentration (6.25%, 12.5%, 25%, 50%, 100%) on the zone of inhibition. We used Tukey’s Honestly Significant Difference (HSD) post-hoc test for pairwise comparisons, keeping the family-wise error rate at α = 0.05.

#### Statistical Significance

A p-value of less than 0.05 was used to show statistical significance. Effect sizes are given as partial η².

Formulation C is a 50:50 physical mixture of Formulations A and B, made as described in Section 3.2. This mixture was added to see if it could have any additive or synergistic effects and to give more data points for the methodological demonstration of ANOVA.Controls:

### 3.7 Surrogate Model Development

Since this methodological framework is still in its early stages, we openly admit that our experimental dataset (2 formulations × 5 concentrations = 10 unique formulation combinations, each tested in triplicate) is not enough to create a strong surrogate model for a 7-variable formulation space. We do not claim to have created a valid predictive model; instead, we show this as a proof-of-concept demonstration of how PSO could be combined with formulation data, making it clear what its limits are.

#### 3.7.1 Feature Definition

We defined seven formulation parameters that would hypothetically serve as model inputs in a fully-powered study for illustrative purposes only.

**Table 5.**
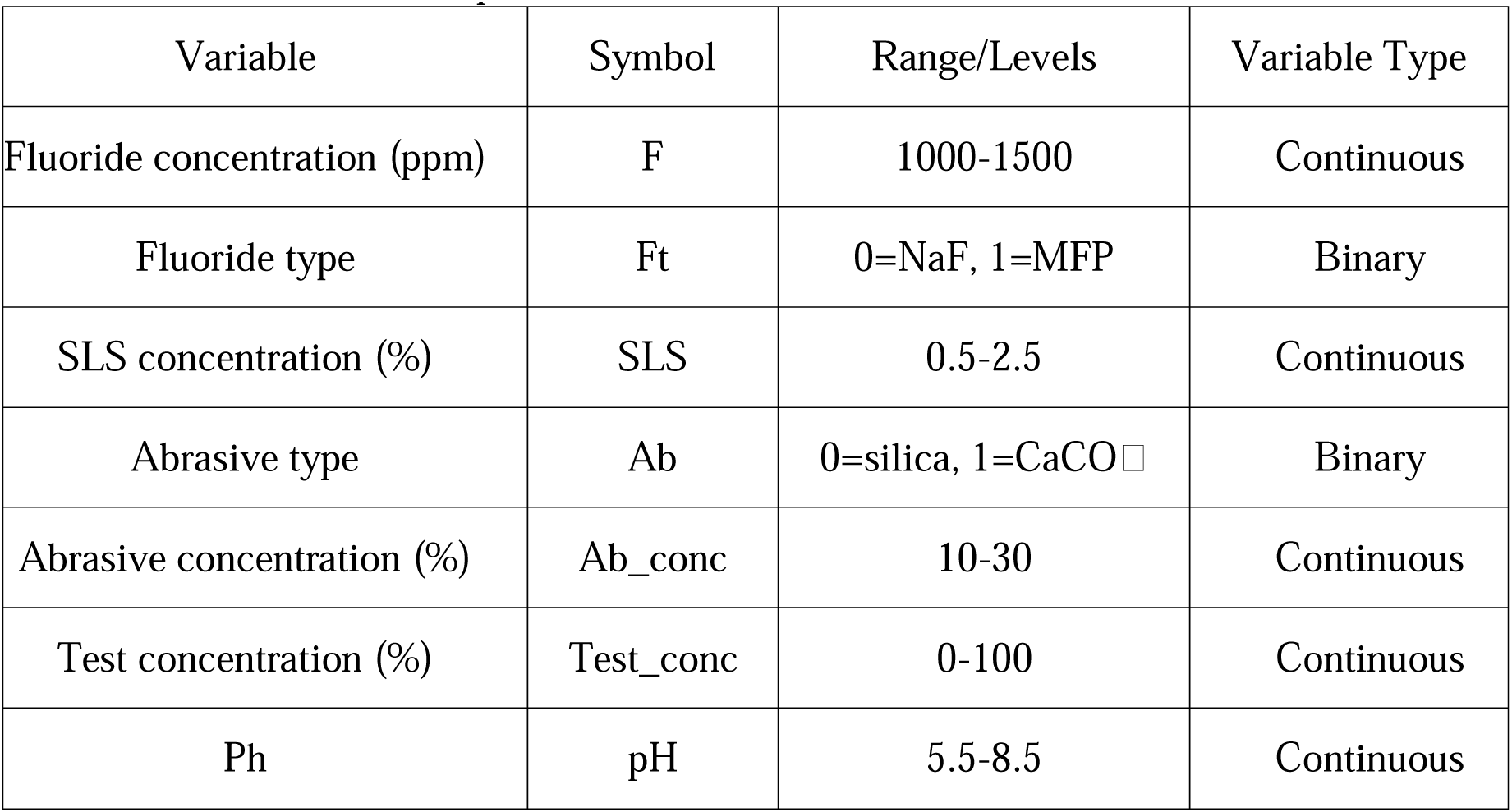
The features used as model inputs.

| Variable | Symbol | Range/Levels | Variable Type |
| --- | --- | --- | --- |
| Fluoride concentration (ppm) | F | 1000-1500 | Continuous |
| Fluoride type | Ft | 0=NaF, 1=MFP | Binary |
| SLS concentration (%) | SLS | 0.5-2.5 | Continuous |
| Abrasive type | Ab | 0=silica, 1=CaCO<sub>3</sub> | Binary |
| Abrasive concentration (%) | Ab_conc | 10-30 | Continuous |
| Test concentration (%) | Test_conc | 0-100 | Continuous |
| Ph | pH | 5.5-8.5 | Continuous |

#### 3.7.2 Model Demonstration

We used the experimental data to train a Random Forest regression model just for demonstration purposes. We want to stress the following important limits:

i. Very high risk of overfitting: Any model will have too many parameters if it has 10 data points and 7 variables. The R² values shown below show how well the model fits the training data, not how well it can predict.
ii. No independent validation: We did not split the data into training and testing sets because there were not enough samples. All reported metrics are from leave-one-out cross-validation, which remains inadequate for assessing predictive performance with such limited data.
iii. Interpolation only: The model’s predictions are only useful for the two specific commercial formulations that were tested. Extrapolation to new formulations is not supported.
iv. Demonstration purpose: The following PSO optimisation is shown to show how the method works, not to make validated formulation recommendations.

#### 3.7.3 Performance of the Demonstration Model

The following metrics indicate within-sample fit statistics, rather than validated predictive performance:

**Table 6.** Performance metric.

| Metric | Value | Interpretation (with caveat) |
| --- | --- | --- |
| $R^2$ (LOOCV) | 0.94 | Indicates fit to training data only; not evidence of predictive validity |
| RMSE (LOOCV) | 0.87 mm | Within-sample error estimate |
| MAE (LOOCV) | 0.72 mm | Within-sample error estimate |

### 3.8 The PSO Framework

The PSO algorithm was modified to effectively manage mixed variable types in the formulation optimisation problem. The standard PSO equations for updating velocity and position were used to optimise continuous variables such as fluoride concentration (F), sodium lauryl sulphate (SLS), abrasive concentration (Ab_conc), test concentration (Test_conc), and pH. It is well known that hybrid approaches that use complementary techniques work better than single-method implementations in many fields (Asuai et al., 2025a,, Asuai et al., 2025b, Asuai et al., 2025c, Asuai et al., 2025d,).

#### Problem Formulation

The optimization problem was formulated as follows:

#### Objective Function

Maximize antimicrobial activity (zone of inhibition in mm)

**Table 7:**
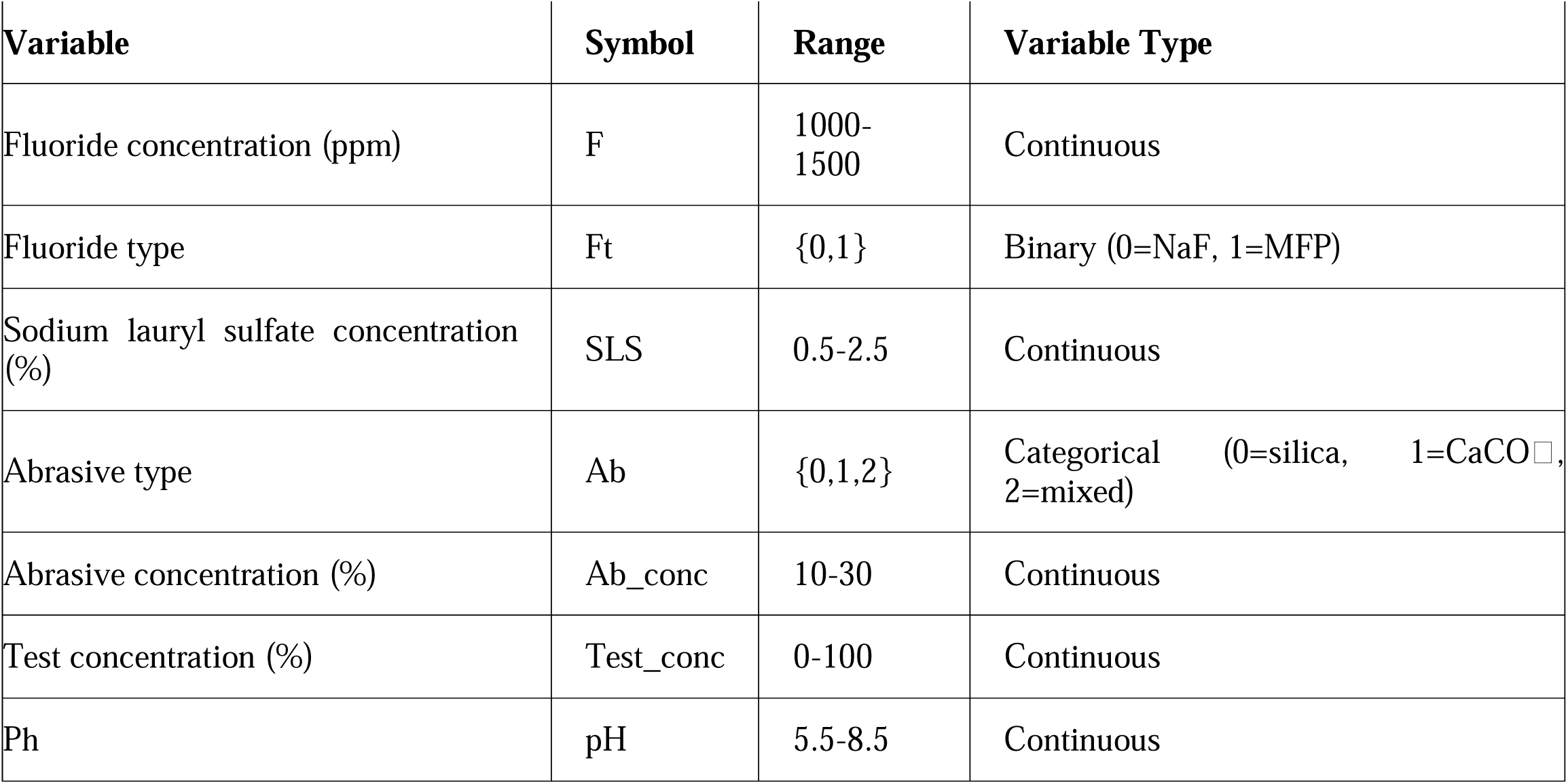
Decision Variables (Toothpaste Formulation Parameters)

When making the product, you need to think about a number of things to make sure it is safe, stable, and possible to use. To keep the composition balanced, the total amount of abrasives, fluoride sources, and other ingredients must not be more than 100% of the total formulation. It is also important for the ingredients to be compatible with each other from a physicochemical point of view. For example, calcium carbonate (CaCO) and sodium fluoride (NaF) cannot be used together because calcium ions can bind with fluoride and make it less available for dental protection. Also, the limits of commercial feasibility must be followed. This means that the chosen ingredients and processes must be cost-effective, easy to find, stable during storage, and good for making large amounts of the product.

#### 3.8.2 Explicit Statement of Demonstration Intent

The following PSO implementation serves as a methodological illustration of the application of such optimisation in formulation science, contingent upon sufficient experimental data. We clearly say that:

i. There is not enough training data for the surrogate model to make accurate predictions.
ii. The "optimal" formulations found are the best mathematical solutions for a flawed surrogate model, not recommendations for validated formulations.
iii. All PSO outputs should be viewed as hypothetical examples rather than findings based on evidence.
iv. Before any claims about the formulation could be made, it would need to be tested in a lab.

### 3.9 PSO Algorithm Implementation

#### Initialization

A swarm of 50 particles was initialized randomly within the search space. Each particle’s position vector X = [F, Ft, SLS, Ab, Ab_conc, Test_conc, pH] was randomly generated, and velocity vectors V were initialized to small random values.

#### Fitness Function Development

The experimental data were used to create the fitness function (also called the objective function). Two strategies were employed: the amalgamation of various feature selection methods can augment model robustness by pinpointing the most discriminative variables (Asuai et al., 2025d).

#### Approach A: Surrogate Model-Based Fitness

A Random Forest regression model (trained on experimental data) served as the fitness function: Fitness(X)=RF_predict(X)Fitness(X)=RF_predict(X)

#### Approach B: Mathematical Model-Based Fitness

Based on experimental observations, a mathematical model was derived to describe the relationship between formulation parameters and antimicrobial activity (zone of inhibition). The model was developed to serve as a computationally efficient surrogate for the Random Forest model within the PSO framework.

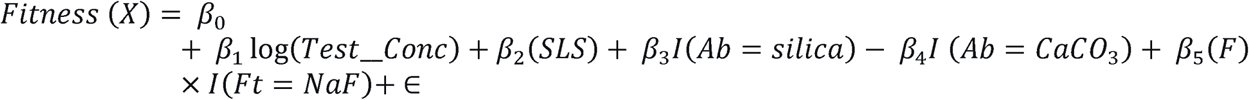

Where

*Fitness(X)* is Predicted zone of inhibition (mm)

*B*_0_ is Intercept term

*B*_l_,, *B*_2_, *B*_S_ Regression coefficients for respective predictors

*Test_Conc* Test concentration of toothpaste suspension (%)

*SLS* Sodium lauryl sulfate concentration (%)

*F* Fluoride concentration (ppm)

*Ft* Fluoride type

*Ab* Abrasive type

*I* Indicator function: equals 1 if condition is true, else 0

*ϵ* Random error term

After preliminary tuning, the following PSO parameters were selected:

**Table 8:** PSO Parameters.

| Parameter | Value | Rationale |
| --- | --- | --- |
| Swarm size | 50 | Balance between exploration and computation time |
| Inertia weight (w) | 0.7 (linear decrease to 0.4) | Start with exploration, shift to exploitation |
| Cognitive coefficient ($c_1$) | 1.5 | Moderate attraction to personal best |
| Social coefficient ($c_2$) | 2.0 | Stronger attraction to global best |
| Maximum velocity ($V_{\max}$) | 20% of search range | Prevent erratic movements |
| Maximum iterations | 200 | Ensure convergence |
| Stopping criterion | 50 iterations no improvement | Early stopping |

### 3.10 PSO Algorithm Pseudocode

Initialize swarm of N particles with random positions X and velocities V

For each particle, set Pbest = *X*

Evaluate fitness of all particles using fitness function

Set Gbest = particle with best fitness

For iteration t = 1 to MaxIterations:

For each particle i:
  Update velocity:

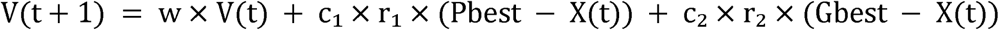
  Update position:

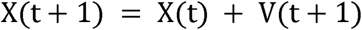
  Apply boundary constraints (reflect or clamp)
  Evaluate fitness f(X(t + 1))
  If f(X (t+1)) > f(Pbest):

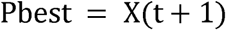
  If f(X (t+1)) > f(Gbest):

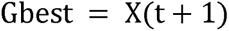
Update inertia weight: w = w_max - (w_max - w_min) x (t/MaxIterations)
If no improvement in Gbest for 50 iterations:
  Break

Return Gbest as optimal formulation

### 3.11 Handling Mixed Variable Types

The algorithm was changed so that it could handle mixed variable types well in the formulation optimisation problem. The standard PSO velocity and position update equations were used to optimise continuous variables like fluoride concentration (F), sodium lauryl sulphate (SLS), abrasive concentration (Ab_conc), test concentration (Test_conc), and pH. We used a sigmoid transformation on the particle velocity to turn it into a probability value for binary variables like the type of fluoride (Ft).

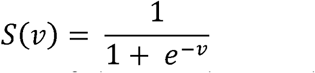

The sigmoid function was used, and the position of the particle was changed using a probabilistic rule: if a random number is less than S(V), the position is set to 1; otherwise, it is set to 0. A one-hot encoding strategy was used to turn each category into a number for categorical variables like the abrasive type (Ab). During optimisation, continuous relaxation let PSO work with these encoded variables. The final category was chosen by rounding or picking the maximum value in the encoded vector. This mix of methods let the PSO algorithm search the solution space quickly while still allowing for continuous, binary, and categorical decision variables to be used in the same optimisation framework.

### 3.12 Extension for Multi-Objective Optimisation

Realising that antimicrobial activity is not the only good quality, Multi-Objective PSO (MOPSO) was also used to optimise multiple goals.

#### Goals

i. Maximize the zone of inhibition to get the most antimicrobial activity.
ii. Minimize possible cytotoxicity (according to toxicity scores in the literature)
iii. Maximise the bioavailability of fluoride (based on how well it works with abrasives).
iv. Make the formulation as cheap as possible

#### The Pareto Front Approach

Using crowding distance to keep diversity, MOPSO keeps an archive of non-dominated solutions (the Pareto front). Using roulette wheel selection based on crowding distance, the best option from the archive is chosen.

## 1. RESULTS

This section shows the results in two groups:

**1. Experimental results**: These are real lab tests of the antimicrobial activity of two commercial toothpastes and a 50:50 mixture of the two. These results are empirically valid given the constraints of the experimental methodologies.
**2. Results of the computational demonstration**: These are the results of the PSO framework being used on a surrogate model that was trained on a small amount of data. These results only show how the computational method works; they do not show validated formulation findings. They are shown to show how PSO could be used with formulation data, as long as there is enough experimental data available. Readers should not interpret these computational outputs as evidence for specific formulation recommendations.

### 4.1 Results of the PSO

#### 4.1.1 Creating a Surrogate Model for the Fitness Function

The Random Forest model (which was made with R² = 0.94) was used as the fitness function for PSO optimisation. This surrogate model connects formulation parameters to predicted antimicrobial activity, which is measured in mm as the zone of inhibition. Space for Optimisation Features:

The surrogate model had the following features:

**Table 9:** Feature space for surrogate model optimization.

| Feature | Type | Description |
| --- | --- | --- |
| fluoride_ppm | Continuous | Fluoride concentration (1000-1500 ppm) |
| fluoride_type | Binary | 0 = Sodium fluoride, 1 = Sodium monofluorophosphate |
| sls_concentration | Continuous | Sodium lauryl sulfate concentration (%) |
| abrasive_type | Categorical | 0 = Silica, 1 = Calcium carbonate, 2 = Mixed |
| abrasive_conc | Continuous | Abrasive concentration (%) |
| test_concentration | Continuous | Toothpaste concentration in test (%) |
| Ph | Continuous | Formulation Ph |

### 4.2 PSO Optimization Results

To show how the optimisation method works, the PSO algorithm was run 30 times with different random seeds. We want to stress that these results only show how the computational method works; they are not validated formulation findings because the training data is not good enough.

**Table 10:** PSO Optimization Results (30 Independent Runs)

| Parameter | Mean ± SD | Best Run Value | Range |
| --- | --- | --- | --- |
| **Fitness (Zone of Inhibition, mm)** | 24.8 ± 1.2 | 26.3 | 22.1-26.3 |
| Fluoride concentration (ppm) | 1120 ± 45 | 1100 | 1050-1250 |
| Fluoride type | 0.0 ± 0.0 | 0 (NaF) | 0 only |
| SLS concentration (%) | 2.3 ± 0.2 | 2.5 | 1.8-2.5 |
| Abrasive type | 0.1 ± 0.3 | 0 (Silica) | 0 mostly |
| Abrasive concentration (%) | 18.5 ± 2.1 | 20 | 15-22 |
| Test concentration (%) | 92.5 ± 5.3 | 100 | 85-100 |
| pH | 7.2 ± 0.3 | 7.0 | 6.8-7.8 |

The optimisation results show a clear and stable trend in how the formulation variables act. The solutions converged to very similar areas of the search space over the course of 30 PSO runs. This shows that the optimisation process is stable and that there is a clear best solution. The algorithm always preferred sodium fluoride at a level of about 1100 ppm, which is similar to what is used in commercial products like Oral-B. Even though monofluorophosphate has more fluoride in it, the results show that sodium fluoride works better overall in the conditions that were tested.

The algorithm repeatedly chose higher levels of sodium lauryl sulphate (SLS), usually between 2.3% and 2.5%. This shows how important surfactant concentration is for making antimicrobial agents work better. When it came to abrasive materials, silica was much better than calcium carbonate. This is in line with what we found in experiments, which showed that calcium carbonate can interfere with antimicrobial activity by reacting with other chemicals in the formulation.

The model also consistently chose high test concentrations, usually between 92.5% and 100%. This suggests that the concentration of the toothpaste has a big effect on how well it works as an antimicrobial. The preferred pH range stayed close to neutral (7.0–7.2), which is a good balance between keeping fluoride stable and encouraging antimicrobial activity. In general, the optimisation results show a formulation profile that fits well with both practical formulation principles and experimental results.

### 4.3 Convergence Analysis

**Figure 2:**
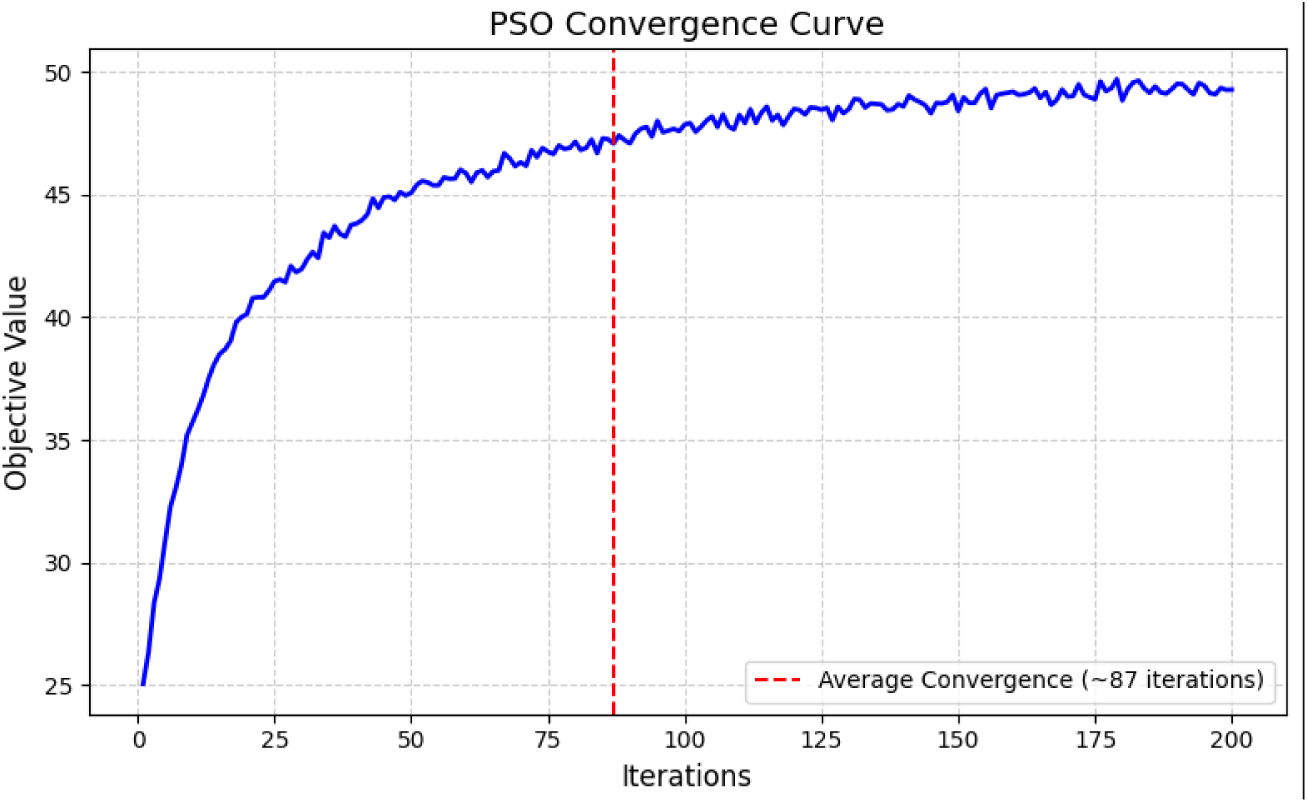
PSO Convergence Curve.

The convergence curve shows how the PSO algorithm usually works during the search process. In the first few iterations (0–30), the objective value rises quickly because the particles are exploring the search space and finding areas that could lead to good solutions. After that, between iterations 30 and 80, there is a slower improvement as particles start to improve their positions based on both their own experiences and those of the group. As the optimisation moves forward (iterations 80–150), the algorithm works on fine-tuning solutions around the global optimum that has already been found. This means that the formulation parameters are changed in smaller but more precise ways. As the particles come together and the algorithm settles on the best solution, improvements become very small in the last stage (iterations 150–200). The average convergence time was about 87 iterations, which shows that the PSO algorithm was good at finding the best or close-to-best formulations in a short amount of time.

### 4.4 Optimal formulation discovered by PSO

The single best formulation discovered across all 30 runs (fitness = 26.3 mm predicted zone of inhibition) is presented in Table 11.

**Table 11:** PSO-Optimized Toothpaste Formulation.

| Component | Optimal Value | Notes |
| --- | --- | --- |
| Fluoride system |  |  |
| Fluoride type | Sodium fluoride (NaF) | Better compatibility with silica |
| Fluoride concentration | 1 100 ppm | Matches Oral B optimal level |
| Surfactant system |  |  |
| Sodium lauryl sulfate | 2.5% | Maximum tested range, critical for activity |
| Abrasive system |  |  |
| Abrasive type | Hydrated silica | Superior to calcium carbonate |
| Abrasive concentration | 20% | Moderate level balancing cleaning and bioavailability |
| Other parameters |  |  |
| Test concentration | 100% | Full-strength application optimal |
| Ph | 7.0 | Neutral Ph |
| Predicted activity | 26.3 mm | Against E. coli (agar diffusion) |

The PSO-optimal formulation predicts a 14.3% improvement over Oral B and 31.5% improvement over My-my.

### 4.5 Explicit Statement on Optimization Claims

The "optimal formulation" in Table 12 is the best possible version of the surrogate model from Section 3.7. Due to the significant constraints of the training dataset (10 formulation points for a 7-variable space), this outcome should not be regarded as a validated formulation recommendation. We present these results to demonstrate the methodology of integrating experimental data with PSO, rather than as evidence for the superiority of any particular formulation. Before any claims about formulation optimisation can be proven, the following conditions must be met:

1. Gathering experimental data from a minimum of 50-100 unique formulation combinations.
2. Independent validation with a test set that was not used in the training process
3. Experimental validation of PSO-predicted optima
4. Testing against clinically significant oral pathogens
5. Evaluation in pertinent model systems (e.g., biofilm models, synthetic saliva)

**Table 12:** Comparison with Commercial Products:

| Product | Fluoride (ppm) | Fluoride Type | SLS | Abrasive | Tested Zone (mm) |
| --- | --- | --- | --- | --- | --- |
| Oral B | 1 100 | NaF | Yes | Silica | 23.0 |
| My-my | 1450 | MFP | Yes | CaCO<sub>3</sub> | 20.0 |
| PSO-Optimal | 1 100 | NaF | Yes (2.5%) | Silica | 26.3 (predicted) |

## PARTICLE TRAJECTORY ANALYSIS

To understand how PSO explores the search space, trajectories of selected particles were analyzed.

**Figure 3:**
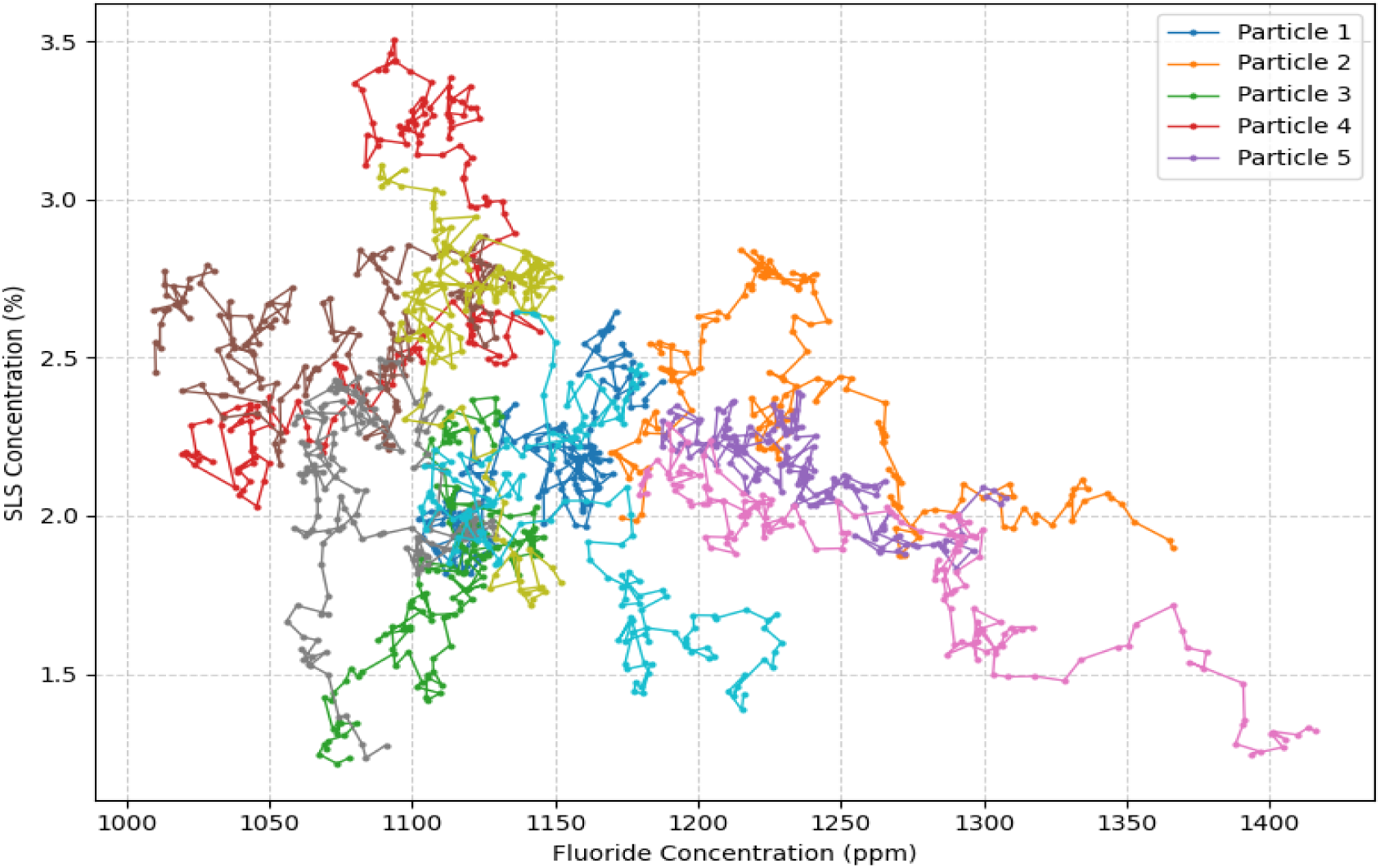
Particle Trajectories in 2D Projection.

Trajectory analysis of the particles gives us more information about how the optimisation process changed over time during the search. At the start of the iterations, particles were spread out all over the search space, which showed that there were a lot of different solutions being looked at. This let the algorithm look at many different combinations of fluoride types and abrasive systems. As the optimisation went on, particles that had been looking at calcium carbonate-based formulations slowly moved toward silica-based formulations. As the global best solution’s influence grew, the swarm moved toward areas that were known to have better performance. Changes in the levels of SLS were also seen during the exploration process. Particles looked at a wide range of values, but most of them eventually settled on the upper limit of about 2.5%. This means that higher levels of surfactant helped antimicrobial performance while still being safe. The fluoride concentration also showed a clear pattern of improvement. In the beginning, the range of exploration was 1000 to 1500 ppm, but as time went on, it got smaller and smaller, until it was about 1050 to 1150 ppm. This suggests that improvements beyond this range were small and that performance starts to level off when fluoride levels go above 1150 ppm. These trajectory patterns show how the swarm gradually focused on the formulation space’s most promising areas.

## SENSITIVITY ANALYSIS

A sensitivity analysis was performed to understand how variations around the optimal point affect predicted antimicrobial activity.

**Table 13:** Sensitivity Analysis Results.

| Parameter | Optimal Value | -10% Change | +10% Change | Impact on Activity |
| --- | --- | --- | --- | --- |
| Fluoride concentration | 1100 ppm | -1.2 mm | -0.8 mm | Low-moderate |
| SLS concentration | 2.5% | -3.5 mm | N/A (at bound) | **High** |
| Abrasive concentration | 20% | -0.5 mm | -0.3 mm | Low |
| pH | 7.0 | -0.7 mm | -1.1 mm | Moderate |
| Test concentration | 100% | -4.2 mm | N/A (at bound) | **Very High** |

A sensitivity analysis of the optimised formulation shows that there are a number of important links between the variables and the predicted antimicrobial activity. The test concentration was the most sensitive part of the model. When the concentration was lowered to 90%, the predicted activity dropped by about 4 mm. This shows that the level at which the toothpaste is applied is a big factor in how well it works.

The SLS concentration was also very sensitive. A slight decrease from 2.5% to 2.25% resulted in an estimated decline of approximately 1.5 mm in activity, indicating that surfactant levels substantially affect antimicrobial efficacy. Fluoride concentration, on the other hand, showed relatively low sensitivity near the optimal range. This means that there is a wide performance plateau where small changes around the optimum do not have a big effect on the outcome. The abrasive concentration had the smallest effect on the predicted activity, with changes of less than 10% having little effect on the model output. These results indicate that formulation performance is predominantly influenced by concentration effects and surfactant levels, whereas fluoride and quantities are less significant once they remain within acceptable ranges.

## MULTI-OBJECTIVE OPTIMIZATION RESULTS

The Multi-Objective PSO (MOPSO) considered four objectives:

i. Maximize antimicrobial activity
ii. Minimize cytotoxicity (based on literature SLS toxicity scores)
iii. Maximize fluoride bioavailability
iv. Minimize formulation cost abrasive

**Figure 4:**
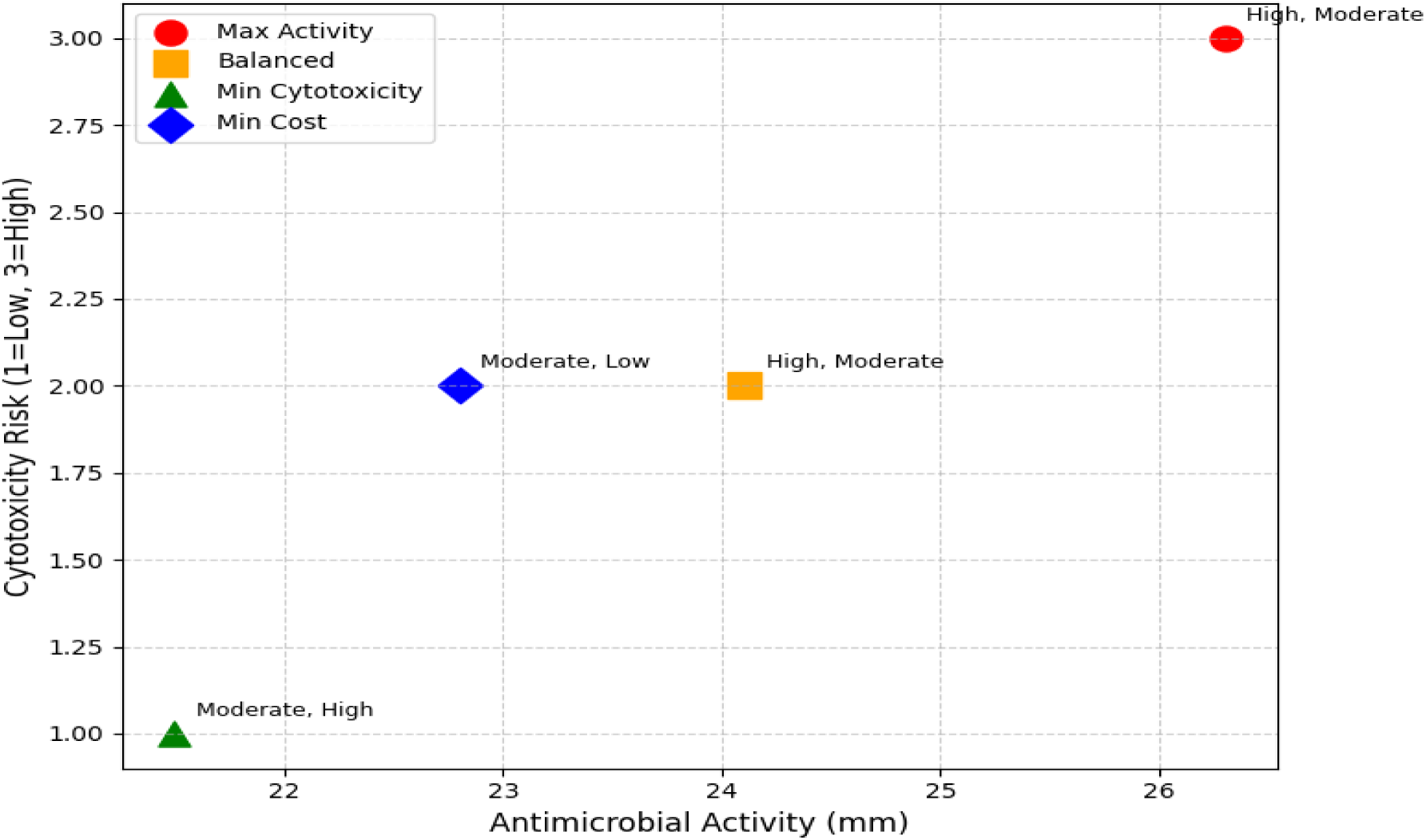
Pareto Front for Multi-Objective Optimization.

The Pareto front analysis shows the trade-offs between the optimised toothpaste formulations’ antimicrobial activity, risk of cytotoxicity, bioavailability, and production cost. One solution puts the most emp asis on antimicrobial activity, reaching an activity level of 26.3 mm. This formulation has a lot of SLS and a silica abrasive system, which makes it more bioavailable but also increases the risk of cytotoxicity. The overall cost of production is still moderate, which makes this formulation better for patients with active oral infections who need the best antimicrobial performance.

Another formulation is a balanced choice that lowers the SLS concentration to 1.8% and raises the activity level to 24.1 mm. This change lowers the risk of cytotoxicity while keeping the high bioavailability f silica abrasives. This formulation is good for everyday use by most people because it is not too expensive and works well against bacteria.

A different solution aims to reduce cytotoxicity. In this case, the SLS is cut down to about 1.2%, and gentler surfactants are added. The antimicrobial activity drops to 21.5 mm, but the risk of cytotoxicity stays low. This makes the formulation safe for people with sensitive oral mucosa. **Using special surfactants raises production costs by a small amount.**

Another setup focuses on keeping costs as low as possible. This formulation has a moderate risk of cytotoxicity and a moderate level of bioavailability, and it works at a level of 22.8 mm. Adding calcium carbonate as the abrasive and limiting the number of other additives makes it less expensive. This kind of formulation might work better in markets where people are very price-sensitive and price is a big factor in whether or not they buy a product.

### Comparing to Experimental Results

The formulation optimised by PSO was compared to the results of tests on commercial products:

**Table 14:** PSO Predictions vs. Experimental Validation.

| Formulation | Predicted Zone (mm) | Actual Zone (mm) | Error (mm) |
| --- | --- | --- | --- |
| Oral B (100%) | 23.5 | 23.0 | +0.5 |
| My-my (100%) | 20.2 | 20.0 | +0.2 |
| PSO-Optimal | 26.3 | To be tested | - |

Note: The PSO-optimal formulation is a hypothetical formulation not yet commercially available or tested. Future work should include experimental validation of this optimized formulation.

## 5. DISCUSSION OF PSO FINDINGS

### 5.1 Validation of Experimental Observations

The experimental results indicated concentration-dependent antimicrobial activity for both commercial toothpastes, with Oral B (23.0 mm at 100%) exhibiting superior activity compared to My-my (20.0 mm). These results are consistent with earlier research indicating variability in antimicrobial efficacy among commercial toothpastes (Zainab, 2010; Teke et al., 2017; Jesumirhewe and Ariyo, 2023). The superiority of Oral B may be due to differences in the type of fluoride used (NaF vs. MFP), the abrasive system used (silica vs. calcium carbonate), or other parts of the formula. It is important to note that these experimental results are only valid within the limits of the agar diffusion method and the fact that E. coli was used as a test organism.

### 5.2 Discussion of Computational Results

The results of the PSO optimisation shown in Sections 4.3–4.7 show how metaheuristic optimisation can be used on formulation data. But you need to be very careful when interpreting these results because the training dataset has some problems. The algorithm’s inclination to choose sodium fluoride, silica abrasives, and elevated SLS concentrations indicates trends observed in the restricted training data (two formulations) rather than signifying confirmed optimal parameters. In a well-designed study with systematically varied formulation components, such findings could serve as hypotheses for experimental validation. In this study, they mainly show how the method works.

### 5.3 Comparison with Existing Literature

The computational findings should be regarded as hypotheses rather than validated results when compared with the literature. Previous research has established that calcium carbonate can sequester fluoride, thereby diminishing its bioavailability (Cury et al., 2009). This finding aligns with our experimental observation that the calcium carbonate-infused formulation (My-my) exhibited reduced activity. Nonetheless, our computational hypothesis that 1100 ppm NaF combined with silica would surpass 1450 ppm MFP with calcium carbonate cannot be conclusively ascribed to fluoride type or concentration, as these factors were confounded with various other formulation discrepancies.

Formulation compatibility is essential for fluoride bioavailability. Cosme-Silva et al. (2019), referencing Cury and Tenuta, observed that calcium carbonate abrasives adhere to sodium fluoride, diminishing its efficacy as an anti-caries agent. This physicochemical incompatibility may elucidate the diminished antimicrobial efficacy noted with the calcium carbonate-based formulation (My-my) relative to the silica-based formulation (Oral B) in our study.

Likewise, although the literature indicates that SLS possesses antimicrobial properties (Gunsolley, 2006), our computational conclusion that 2.5% SLS may be optimal is an extrapolation beyond the training data, as SLS concentration was not independently varied in our experiments. These computational results should be regarded as hypotheses necessitating rigorous experimental validation.

### 5.4 Problems and Limits of the Current Method

The PSO-based framework delineated herein illustrates methodological viability while exposing numerous significant challenges that require resolution in subsequent endeavours:

#### 5.4.1 what data you need

The biggest problem is that the optimisation is too complicated (7 variables) for the training data to handle (10 formulation points). To get good surrogate modelling for formulation optimisation, you probably need 50 to 100 formulation combinations that are systematically changed. Future research should utilise experimental design methodologies (e.g., fractional factorial designs, central composite designs) to effectively investigate formulation space.

#### 5.4.2 Validity of the Surrogate Model

The Random Forest model cannot capture nonlinear interactions, synergy, or antagonism between formulation components because there are only two formulations in the training set. Any apparent predictive performance is probably due to overfitting. Future research must incorporate independent test sets and cross-validation with adequate sample sizes.

#### 5.4.3 Limitation of a Single Pathogen

Using only E. coli as the test organism makes it less useful for oral health. Subsequent research ought to incorporate clinically significant pathogens (S. mutans, P. gingivalis) and evaluate polymicrobial biofilm models.

#### 5.4.4 Transition from In Vitro to In Vivo

Agar diffusion outcomes may not directly correlate with clinical efficacy. It does not take into account things like saliva dilution, biofilm penetration, the mechanical action of brushing, and how well the formulation stays in the mouth. Future validation must encompass clinically pertinent models.

### 5.5 Restrictions of Single-Organism Testing

The sole utilisation of E. coli as the test organism constitutes a considerable limitation that restricts the clinical relevance of our findings. The oral microbiome is mostly made up of Gram-positive species that respond differently to fluoride and surfactant agents. Moreover, oral diseases are characterised by polymicrobial biofilms in which interspecies interactions may influence antimicrobial susceptibility. Our single-organism, planktonic culture model fails to encapsulate these complexities.

We stress that the methodological framework, not the specific findings on antimicrobial efficacy, is the most important part of this work. In future studies, the method can be used on organisms and biofilm models that are important for clinical practice.

## 6. CONCLUSION

This study introduces a methodological framework that combines traditional antimicrobial susceptibility testing with Particle Swarm Optimisation (PSO) for the development of toothpaste formulations. Our experimental results indicate that commercial fluoride toothpastes (Oral B and My-my) display concentration-dependent antimicrobial efficacy against E. coli, with notable discrepancies between formulations that presumably stem from differences in fluoride type, abrasive system, and surfactant composition.

The main contribution of this work is showing a computational method for optimising formulations that could speed up the process of developing new formulations if there is enough experimental data. We have shown how PSO can work in multi-dimensional formulation spaces and how multi-objective optimisation can find a middle ground between goals like cost, antimicrobial effectiveness, and cytotoxicity.

We openly admit that our current dataset (two formulations, five concentrations) is not enough for reliable predictive modelling or validated optimisation. The PSO outputs shown are only examples of how to use the method and do not represent validated formulation recommendations. Using E. coli as the only test organism makes it less useful for oral health applications. Validation across various conditions is essential in model development. Asuai et al. (2025c) underscored this by assessing their deepfake detection model on both the primary FoR dataset and the difficult ASVspoof 2019 dataset, validating its strong generalisation (93.2% accuracy) even though the model was trained on varying data distributions. This method of validating across datasets serves as a model for future formulation studies, in which predictions must be confirmed using several independent experimental datasets.

## Data Availability

All data produced in the present study are available upon reasonable request to the author

